# Effect of Stay-at-Home Orders and Other COVID-Related Policies on Trauma Hospitalization Rates and Disparities in the United States: A Statewide Time-Series Analysis

**DOI:** 10.1101/2022.07.11.22277511

**Authors:** Paula D Strassle, Alan C Kinlaw, Jamie S Ko, Stephanie M Quintero, Jackie Bonilla, Madison Ponder, Anna María Nápoles, Sharon E Schiro

## Abstract

**Background:** To combat the coronavirus pandemic, states implemented several public health policies to reduce infection and transmission. Increasing evidence suggests that these prevention strategies also have had a profound impact on non-COVID healthcare utilization. The goal of this study was to determine the impact of a statewide Stay-at-Home and other COVID-related policies on trauma hospitalizations, stratified by race/ethnicity, age, and sex.

**Methods:** We used the North Carolina Trauma Registry, a statewide registry of trauma hospitalizations to 18 hospitals across North Carolina, including all North Carolina trauma centers, to calculate weekly assault, self-inflicted, unintentional motor vehicle collision (MVC), and other unintentional injury hospitalization rates between January 1, 2019 and December 31, 2020. Interrupted time-series design and segmented linear regression were used to estimate changes in hospitalizations rates after several COVID-related executive orders, overall and stratified by race/ethnicity, age, and gender. Hospitalization rates were compared after 1) U.S. declaration of a public health emergency; 2) North Carolina statewide Stay-at-Home order; 3) Stay-at-Home order lifted with restrictions (Phase 2: Safer-at-Home); and 4) further lifting of restrictions (Phase 2.5: Safer-at-Home).

**Results:** There were 70,478 trauma hospitalizations in North Carolina from 2019-2020. In 2020, median age was 53 years old and 59% were male. Assault hospitalization rates (per 1,000,000 NC residents) increased after the Stay-at-Home order, but only among Black/African American residents (incidence rate difference [IRD]=7.9; other racial/ethnic groups’ IRDs ranged 0.9 to 1.7) and 18-44 year-old males (IRD=11.9; other sex/age groups’ IRDs ranged -0.5 to 3.6). After major restrictions were lifted, assault rates returned to pre-COVID levels. Unintentional injury hospitalizations decreased after the public health emergency, especially among older adults, but returned to 2019 levels within several months.

**Conclusions:** Statewide Stay-at-Home orders put Black/African American residents at higher risk for assault hospitalizations, exacerbating pre-existing disparities. Fear of COVID-19 may have also led to decreases in unintentional non-MVC hospitalization rates, particularly among older adults. Policy makers must anticipate possible negative effects and develop approaches for mitigating harms that may disproportionately affect already disadvantaged communities.

## Introduction

To combat the coronavirus pandemic, many countries implemented lockdowns and Stay-at-Home orders in 2020 to reduce transmission(1); in the United States, these policies were implemented on a state-by-state basis(2). While these orders had a relatively positive impact on reducing COVID-19 infections(1-3), increasing evidence suggests that these prevention strategies also have had a profound impact on non-COVID healthcare utilization. For instance, Stay-at-Home orders in the United States have been associated with decreases in emergency department (ED) visits(4, 5), trauma admissions(6, 7), and motor vehicle collisions (MVCs)(6, 8-10) as well as increases in suicide/suicidal attempts(6, 11-13), firearm injuries(6, 9, 14, 15), and domestic violence/child abuse(16-18).

Despite trauma patients having presumed equal access to healthcare and the highly protocolized nature of trauma management plans, racial and ethnic disparities in the United States were prevalent in traumatic injury prior to the COVID-19 pandemic(19, 20). Minority and low income individuals in the U.S. were also more likely to have public-facing occupations that required them to continue to work in person during the pandemic and Stay-at-Home orders(21, 22). Minority and low income individuals are also more likely to have crowded living conditions(22), which may place them at higher risk of domestic violence and other trauma during the pandemic. For these reasons and others, it is possible that Stay-at-Home orders and other COVID-related policies exacerbated known racial/ethnic disparities in trauma and non-COVID related hospitalizations. Gaining a better understanding of the burden and potential exacerbation of traumatic injury disparities in the United States during the pandemic is necessary to inform and appropriately address mitigation efforts and related policies.

Thus, the goal of this study was to assess changes in traumatic injury hospitalization rates during the first year of the pandemic in the U.S. and assess how Stay-at-Home orders or COVID-related policies were associated with these changes. We were also interested in assessing whether the potential impact of these policies were similar across race/ethnicity, age, and sex.

## Methods

We used data from the North Carolina Trauma Registry (NCTR), a statewide registry and cooperative effort between eighteen North Carolina hospitals, including all 17 North Carolina trauma centers (6 Level I, 3 Level II, and 8 Level III hospitals) and the North Carolina Office of Emergency Medical Services (NCOEMS)(23, 24). This registry, which has been in place since 1987, collects near real-time information using standardized data definitions based off of the National Trauma Registry of the American College of Surgeons and designated NCTR chart abstractors(23, 24). The NCTR includes all hospitalizations where a patient is diagnosed with a traumatic injury (ICD-10-CM: S00-S99, T07, T14, T20-T28, T30-T32, T71, T79.A1-T79.A9), and is admitted to the hospital, taken to the operating room from the emergency department, transferred, or dies due to their injury. Unplanned readmissions within 30 days of the initial injury are also included.

For this study, we included all trauma hospitalizations that occurred between January 1, 2019 and December 31, 2020. To account for variation between weekday and weekend hospitalization rates, we calculated the weekly hospitalization rates for traumatic injuries per 1,000,000 North Carolina residents between January 6, 2019, and December 26, 2020. Admissions that occurred during partial weeks (January 1-5, 2019 and December 27-31, 2020) were excluded from modeling to avoid introducing bias due to underestimation (486 and 393 hospitalizations, respectively; in 2019 there were an average of 682 [SD 61.9] trauma hospitalizations per week). North Carolina population counts for 2019 were obtained from the North Carolina Office of State Budget and Management(25) and were used for both 2019 and 2020 hospitalization rate calculations.

Trauma admissions were classified by injury intent and mechanism into four categories – assault, self-inflicted, unintentional MVC (including MVC-bicyclist and MVC-pedestrian injuries), and unintentional non-MVC – using the ICD-10-CM code framework from the National Center for Health Statistics and National Center for Injury Prevention and Control(26).

Data on race and ethnicity were used to categorize hospitalized patients as non-Hispanic Black/African American, Hispanic/Latino, non-Hispanic White, and non-Hispanic other race. Other race included American Indian (n=513 hospitalizations), Asian (n=596 hospitalizations), Pacific Islander (n=87 hospitalizations), multiracial (n=202 hospitalizations), and those who listed “other” race (n=1,048 hospitalizations). Race and ethnicity were self-reported by the patient (or family member) if they were present and capable; otherwise, it was based on staff designation in the electronic medical record.

COVID-related policies of interest included: U.S. declaration of a public health emergency (1/31/2020), the North Carolina statewide Stay-at-Home order (3/30/2020), an initial lifting of the Stay-at-Home order with restrictions (Phase 2: Safer-at-Home, 5/22/2020), and the further lifting of Stay-at-Home restrictions (Phase 2.5: Safer-at-Home, 9/4/2020), Supplemental Table 1. Policies were assigned to the week of their effective date. Other statewide executive orders that were not included in analyses were North Carolina declaring a state of emergency, statewide closure of K-12 public schools, Phase 1 and Phase 3 of lifting the statewide Stay-at-Home orders, and the modified Stay-at-Home order issued before the 2020 holidays. These orders were not included in analyses because either the order made relatively small changes to existing orders (e.g., Phase 1 lifting of Stay-at-Home orders) or it occurred within several weeks of a prior order that we believed would be more salient (e.g., North Carolina declaring a state of emergency).

Differences in patient demographics and clinical characteristics among patients admitted for traumatic injuries between 2019 and 2020 were compared using standardized differences. An absolute difference >0.20 was considered meaningful.

We conducted a natural experiment using an interrupted-time series design and segmented linear regression(27, 28). Using ordinary least squares, we conducted injury intent and mechanism-specific segmented linear regression models to estimate the trend in trauma hospitalization rates between each pair of interruptions. Our models did not include parameters for level changes (i.e., intercept changes) to focus our analysis on *a priori*-hypothesized gradual changes in injury hospitalization rates. To reduce error in our model, we used a transformed cosine periodic function to control for potential seasonal fluctuations in hospitalization rates(29). To account for autocorrelation over time, we used Durbin-Watson tests (α=0.05) to specify autoregressive parameters in our models for lags up to 60 weeks. Similar methods were used to estimate race/ethnicity-specific and age and sex-specific hospitalization rates. Due to low overall rates, race/ethnicity and age/sex-stratified models for self-inflicted injuries were not performed.

All analyses were performed using SAS version 9.4 (SAS Inc., Cary, North Carolina). This study was deemed exempt by the University of North Carolina (IRB# 20-2117) and National Institutes of Health (IRB# 000330) Institutional Review Boards.

## Results

Between 2019 and 2020, there were 70,478 trauma hospitalizations at participating sites (including all trauma centers); 43.6% (n=30,712) occurred after COVID-19 was declared a U.S. public health emergency. In 2020, there were 354 confirmed and 5,543 suspected COVID-19 cases (16.9%) among hospitalized trauma patients. Demographics and clinical characteristics remained relatively consistent between 2019 and 2020, Table 1. The most common types of trauma admissions by intent/mechanism and year were unintentional non-MVC (2019: 60.5%; 2020: 59.6%), followed by unintentional MVCs (2019: 29.0%; 2020: 28.8%), assaults (2019: 9.3%; 2020: 10.4%), and then self-inflicted injuries (1.2% in both years). The majority of unintentional non-MVC hospitalizations between 2019 and 2020 were falls (n=32,092, 77.4%).

**Table 1.**
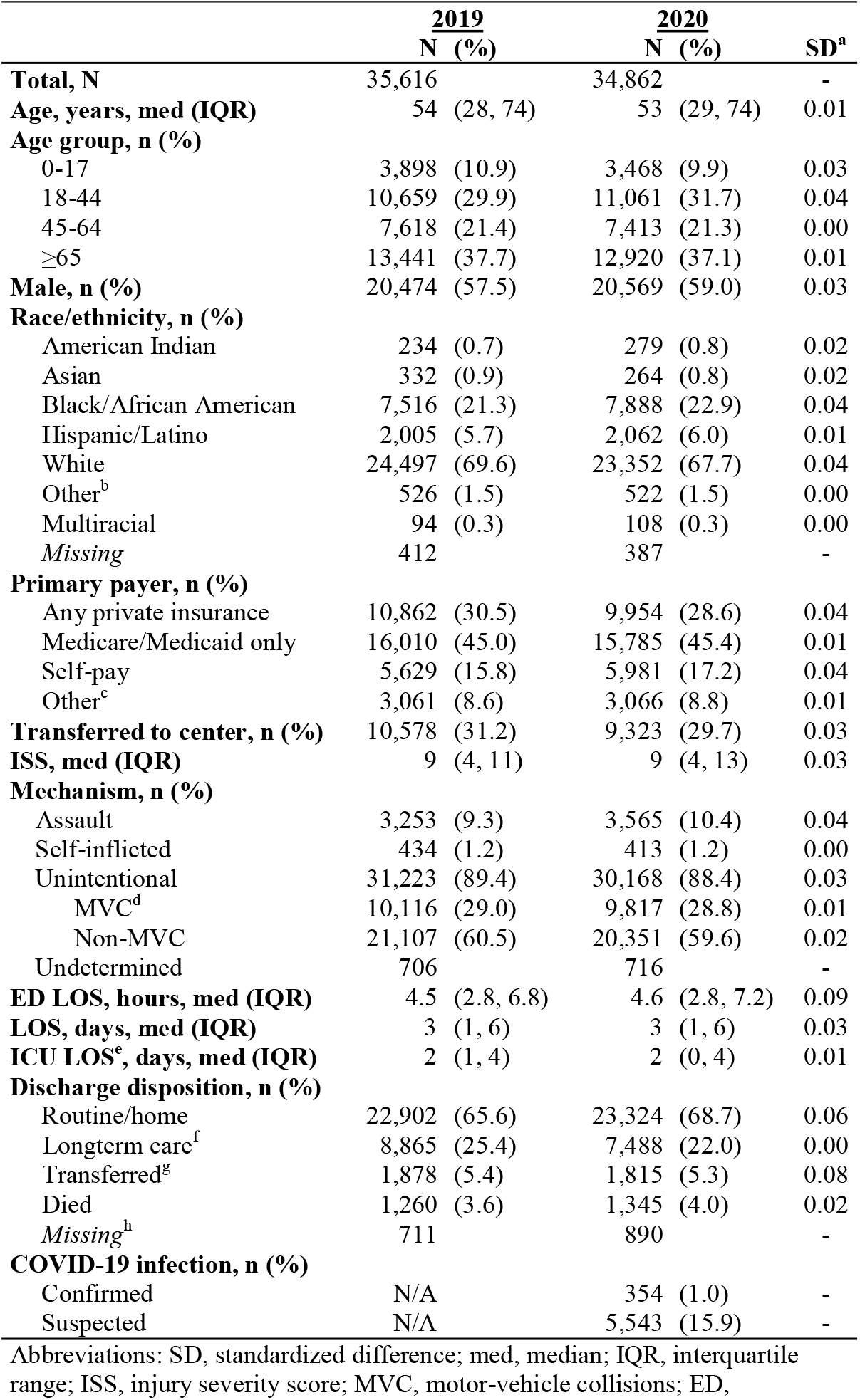
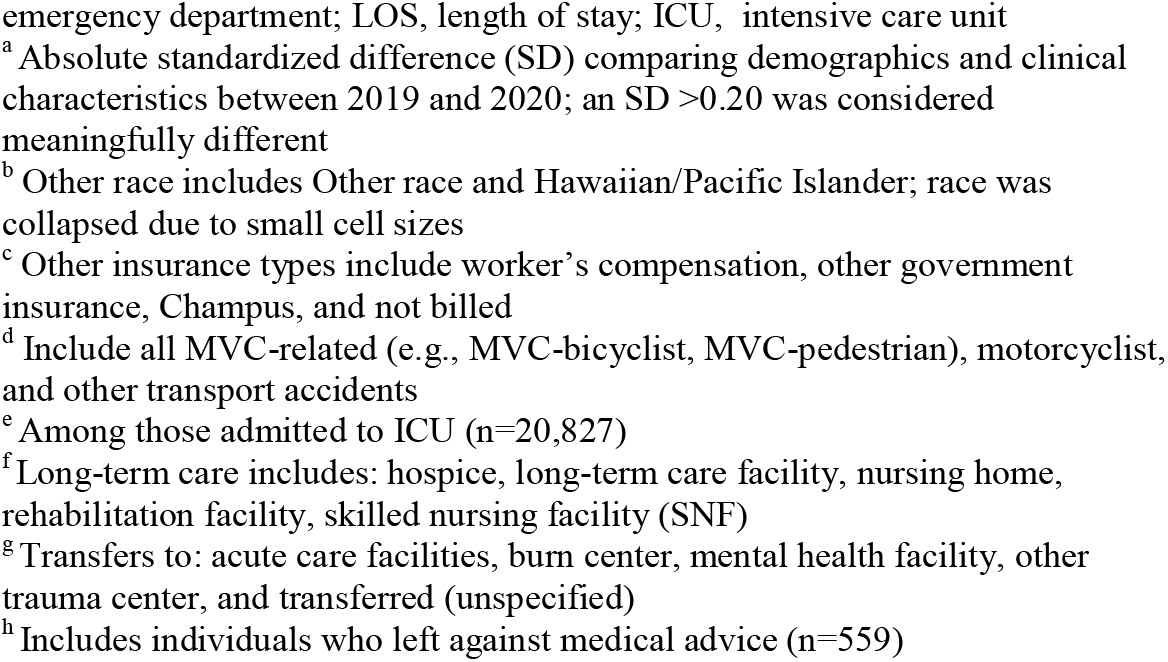
Demographics and clinical characteristics of trauma hospitalizations captured in the North Carolina Trauma Registry between 2019 and 2020, stratified by year.

|  | **2019** | **2020** | **SD<sup>a</sup>** |
| --- | --- | --- | --- |
|  | **N (%)** | **N (%)** |  |
| **Total, N** | 35,616 | 34,862 | - |
| **Age, years, med (IQR)** | 54 (28, 74) | 53 (29, 74) | 0.01 |
| **Age group, n (%)** |  |  |  |
| 0-17 | 3,898 (10.9) | 3,468 (9.9) | 0.03 |
| 18-44 | 10,659 (29.9) | 11,061 (31.7) | 0.04 |
| 45-64 | 7,618 (21.4) | 7,413 (21.3) | 0.00 |
| ≥65 | 13,441 (37.7) | 12,920 (37.1) | 0.01 |
| **Male, n (%)** | 20,474 (57.5) | 20,569 (59.0) | 0.03 |
| **Race/ethnicity, n (%)** |  |  |  |
| American Indian | 234 (0.7) | 279 (0.8) | 0.02 |
| Asian | 332 (0.9) | 264 (0.8) | 0.02 |
| Black/African American | 7,516 (21.3) | 7,888 (22.9) | 0.04 |
| Hispanic/Latino | 2,005 (5.7) | 2,062 (6.0) | 0.01 |
| White | 24,497 (69.6) | 23,352 (67.7) | 0.04 |
| Other<sup>b</sup> | 526 (1.5) | 522 (1.5) | 0.00 |
| Multiracial | 94 (0.3) | 108 (0.3) | 0.00 |
| Missing | 412 | 387 | - |
| **Primary payer, n (%)** |  |  |  |
| Any private insurance | 10,862 (30.5) | 9,954 (28.6) | 0.04 |
| Medicare/Medicaid only | 16,010 (45.0) | 15,785 (45.4) | 0.01 |
| Self-pay | 5,629 (15.8) | 5,981 (17.2) | 0.04 |
| Other<sup>c</sup> | 3,061 (8.6) | 3,066 (8.8) | 0.01 |
| **Transferred to center, n (%)** | 10,578 (31.2) | 9,323 (29.7) | 0.03 |
| **ISS, med (IQR)** | 9 (4, 11) | 9 (4, 13) | 0.03 |
| **Mechanism, n (%)** |  |  |  |
| Assault | 3,253 (9.3) | 3,565 (10.4) | 0.04 |
| Self-inflicted | 434 (1.2) | 413 (1.2) | 0.00 |
| Unintentional | 31,223 (89.4) | 30,168 (88.4) | 0.03 |
| MVC<sup>d</sup> | 10,116 (29.0) | 9,817 (28.8) | 0.01 |
| Non-MVC | 21,107 (60.5) | 20,351 (59.6) | 0.02 |
| Undetermined | 706 | 716 | - |
| **ED LOS, hours, med (IQR)** | 4.5 (2.8, 6.8) | 4.6 (2.8, 7.2) | 0.09 |
| **LOS, days, med (IQR)** | 3 (1, 6) | 3 (1, 6) | 0.03 |
| **ICU LOS<sup>e</sup>, days, med (IQR)** | 2 (1, 4) | 2 (0, 4) | 0.01 |
| **Discharge disposition, n (%)** |  |  |  |
| Routine/home | 22,902 (65.6) | 23,324 (68.7) | 0.06 |
| Longterm care<sup>f</sup> | 8,865 (25.4) | 7,488 (22.0) | 0.00 |
| Transferred<sup>g</sup> | 1,878 (5.4) | 1,815 (5.3) | 0.08 |
| Died | 1,260 (3.6) | 1,345 (4.0) | 0.02 |
| Missing<sup>h</sup> | 711 | 890 | - |
| **COVID-19 infection, n (%)** |  |  |  |
| Confirmed | N/A | 354 (1.0) | - |
| Suspected | N/A | 5,543 (15.9) | - |

Abbreviations: SD, standardized difference; med, median; IQR, interquartile range; ISS, injury severity score; MVC, motor-vehicle collisions; ED,
emergency department; LOS, length of stay; ICU, intensive care unit
<sup>a</sup> Absolute standardized difference (SD) comparing demographics and clinical characteristics between 2019 and 2020; an SD >0.20 was considered meaningfully different
<sup>b</sup> Other race includes Other race and Hawaiian/Pacific Islander; race was collapsed due to small cell sizes
<sup>c</sup> Other insurance types include worker's compensation, other government insurance, Champus, and not billed
<sup>d</sup> Include all MVC-related (e.g., MVC-bicyclist, MVC-pedestrian), motorcyclist, and other transport accidents
<sup>e</sup> Among those admitted to ICU (n=20,827)
<sup>f</sup> Long-term care includes: hospice, long-term care facility, nursing home, rehabilitation facility, skilled nursing facility (SNF)
<sup>g</sup> Transfers to: acute care facilities, burn center, mental health facility, other trauma center, and transferred (unspecified)
<sup>h</sup> Includes individuals who left against medical advice (n=559)

In 2019 the weekly hospitalization rates of intentional (assault and self-inflicted injuries) and unintentional (MVCs and non-MVCs) injuries were stable; however, substantial changes were seen early on in the pandemic, Figure 1 and Supplemental Table 2. After the statewide Stay-at-Home order was issued, the rate of assault hospitalizations increased from 5.8 to 8.0 assault hospitalizations per 1,000,000 North Carolina residents (incident rate difference [IRD]=2.2, incident rate ratio [IRR]=1.38) by the time restrictions began to be lifted with Phase 2: Safer-at-Home. Self-inflicted injury hospitalization rates also increased between these two executive orders from 0.5 to 0.9 hospitalizations per 1,000,000 (IRD=0.4, IRR=1.80). After Stay-at-Home orders were lifted and Safer-at-Home began, which reopened businesses with limited capacity, both overall assault and self-inflicted injury hospitalization rates returned to 2019 levels, Figure 1A.

**Figure 1.**
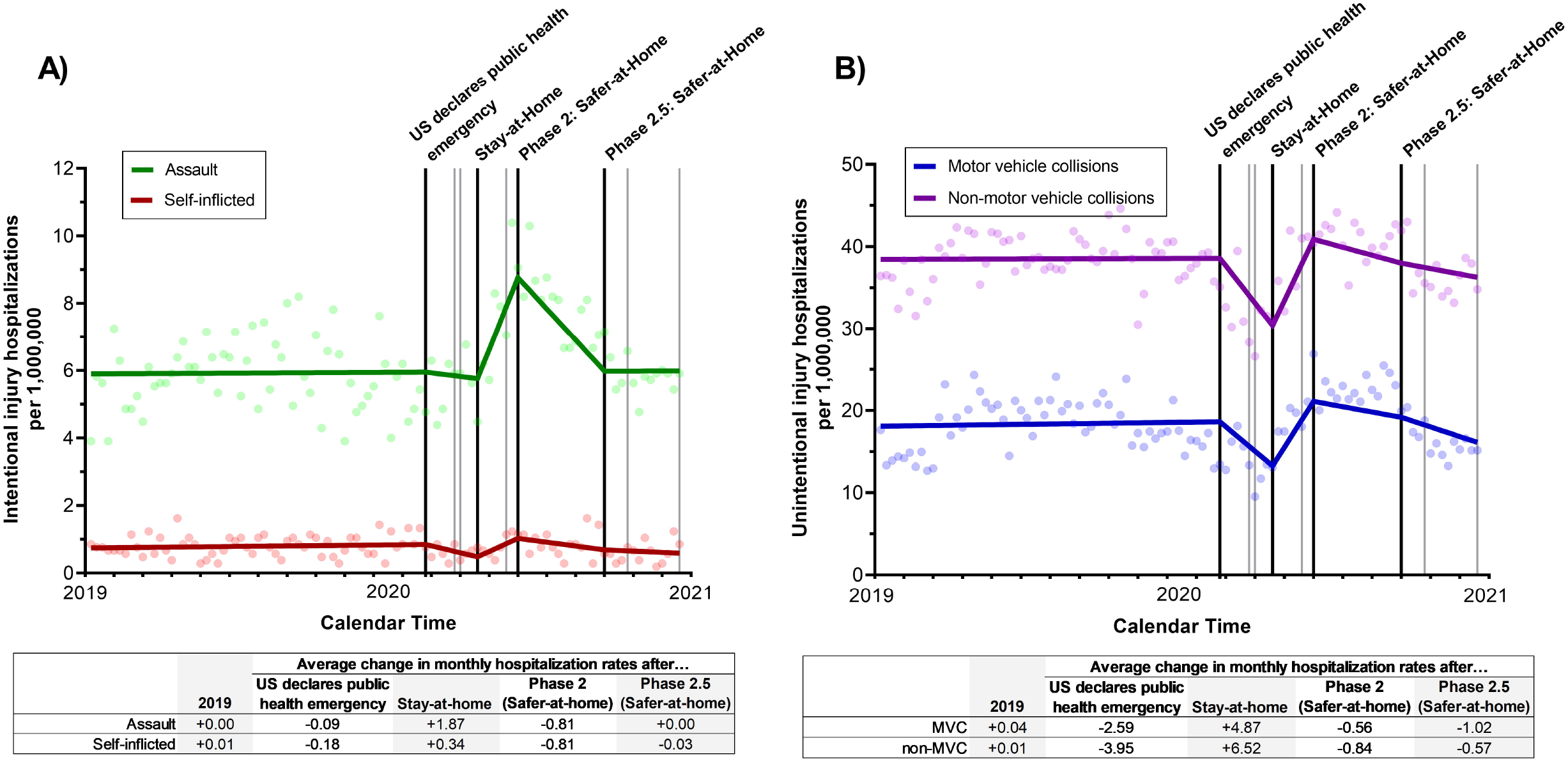
Overall impact of COVID-19 executive orders on weekly number of trauma admissions to trauma centers for A) intentional and B) unintentional injuries between January 2019 and December 2020 in North Carolina. The black lines represent the timing of the four executive orders assessed in the analyses (US declares public health emergency, North Carolina statewide Stay-at-Home order, statewide Phase 2: Safer-at-Home order, and statewide Phase 2.5: Safer-at-Home order); grey lines represent the time of the other COVID-related executive orders.

Both unintentional MVCs and unintentional non-MVC injury hospitalization rates dropped after the U.S. declaration of a public health emergency, with the lowest estimated rates seen at the time Stay-at-Home order was issued (MVC: decreased from 18.6 to 13.3 per 1,000,000 [IRD=-5.3, IRR=0.72]; non-MVC: decreased from 38.6 to 30.4 per 1,000,000 [IRD=-8.4, IRR=0.79]). Unintentional injury (MVC and non-MVC) hospitalization rates began increasing after the Stay-at-Home orders were implemented, and by the end of 2020 MVC and non-MVC injury hospitalization rates had largely returned to 2019 levels, Figure 1B.

### Disparities in assault hospitalization rates during COVID-19

Prior to the COVID-19 pandemic, substantial racial/ethnic disparities were seen in North Carolina assault hospitalization rates; Black/African American North Carolina residents had over 5 times the rate of hospitalizations with assault injuries compared to White, Hispanic/Latino, and other race residents, Figure 2A. Between when the Stay-at-Home order was issued and when major restrictions were first lifted (Phase 2: Safer at Home), this disparity widened due to a sharp absolute increase in assault hospitalization rates among Black/African American North Carolina residents (from 16.7 to 24.6 per 1,000,000, [IRD=7.9, IRR=1.47]), Supplemental Table 2. Minimal changes were seen in assault hospitalization rates among Hispanic/Latino, White, and other race residents during this time (IRDs 0.9-1.8 per 100,000). By the beginning of Phase 2.5: Safer-at-Home assault hospitalization rates among Black/African Americans returned to 2019 levels.

**Figure 2.**
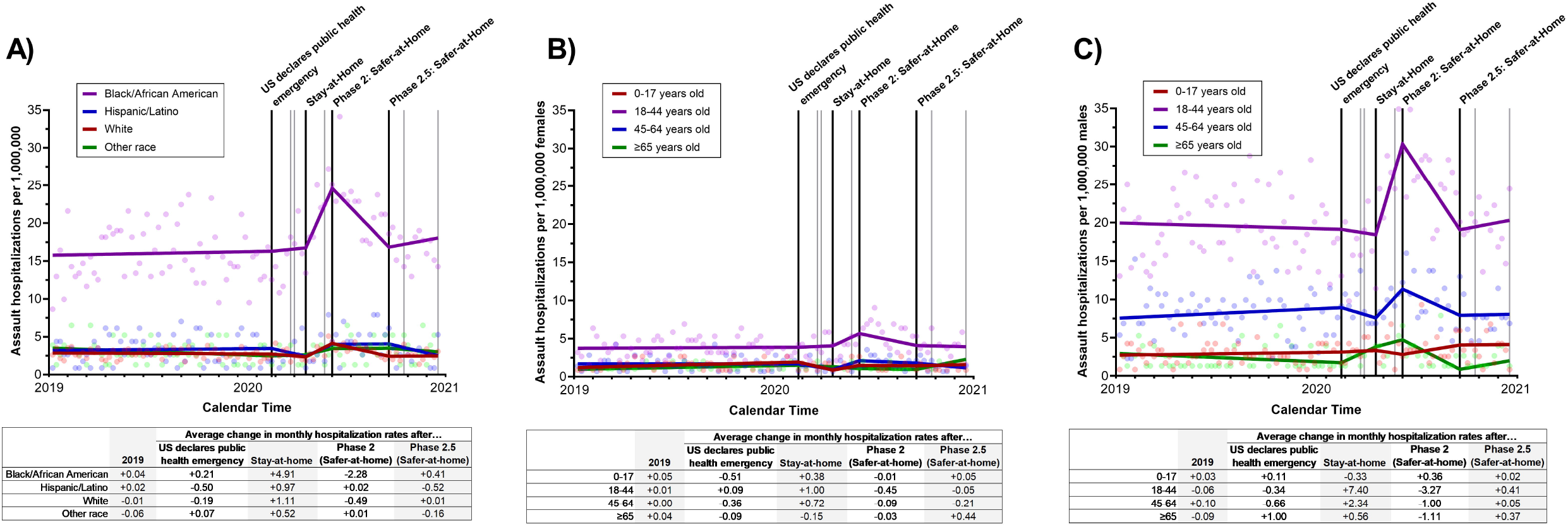
Impact of COVID-19 executive orders on weekly number of assault admissions to trauma centers between January 2019 and December 2020 in North Carolina, stratified by A) race/ethnicity, B) age group among females, and C) age group among males. The black lines represent the timing of the four executive orders assessed in the analyses (US declares public health emergency, North Carolina statewide Stay-at-Home order, statewide Phase 2: Safer-at-Home order, and statewide Phase 2.5: Safer-at-Home order); grey lines represent the time of the other COVID-related executive orders.

Among both males and females, after Stay-at-Home orders were issued, increases in assault hospitalizations were largely observed among young adults (18-44 years old), with almost no changes seen in children or older adults (≥65 years old), Figures 2B and 2C. Assault hospitalization rates among 18-44 year-old males were substantially higher compared to both females (all ages) and their other male counterparts. Among 18-44 year-old males, assault hospitalization rates increased from 18.4 to 30.3 hospitalizations per 1,000,000 males (IRD=11.9, IRR=1.65) between when the Stay-at-Home and Phase 2: Safer-at-Home orders were issued, Figure 2C and Supplemental Table 3. After the Stay-at-Home order was lifted (Phase 2: Safer-at-Home), assault hospitalization rates dropped back to 2019 levels.

### Disparities in unintentional MVC hospitalization rates during COVID-19

Fewer disparities were seen in unintentional MVC hospitalization rates in North Carolina, Figure 3. Across all racial/ethnic and age/sex groups, the rates of unintentional MVC hospitalizations dropped after the U.S. declared a public health emergency (Figure 3A, Supplemental Table 2), but began to return to 2019 levels after a few months. Black/African American residents experienced a more substantial spike in unintentional MVC hospitalizations compared to other racial/ethnic groups (average monthly increase of 8.27 hospitalizations per 1,000,000 compared to 3.84-5.13 in other racial/ethnic groups) and did not fall back to 2019 rates until the end of 2020.

**Figure 3.**
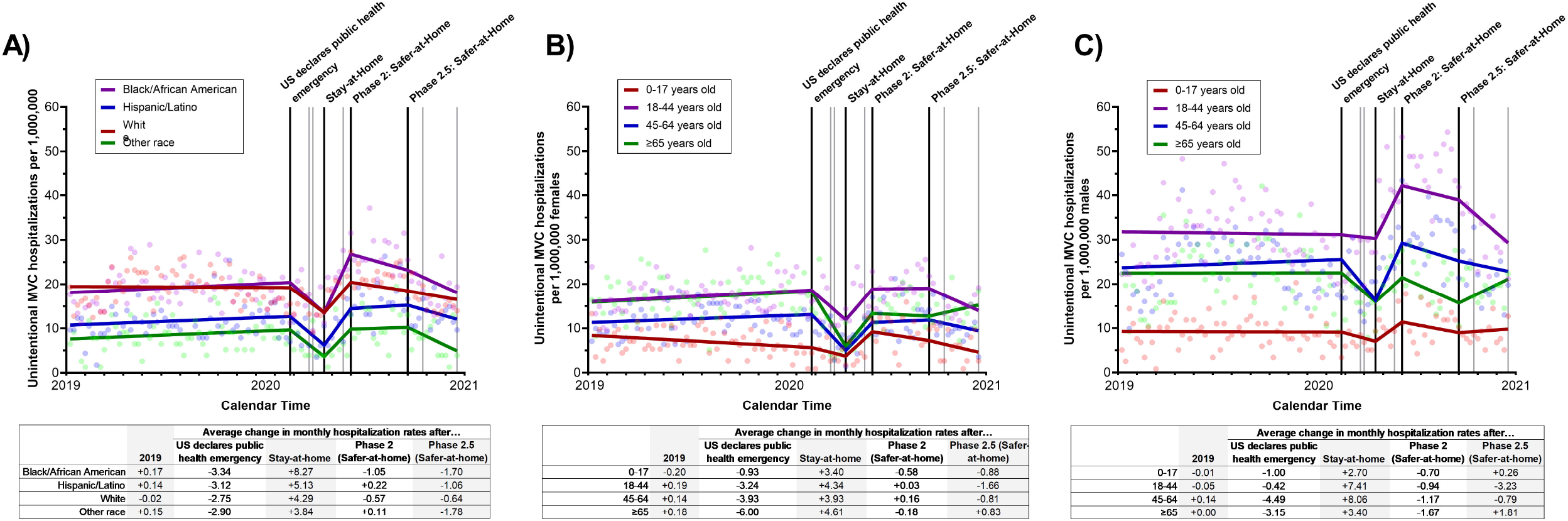
Impact of COVID-19 executive orders on weekly number of unintentional MVC admissions to trauma centers between January 2019 and December 2020 in North Carolina, stratified by A) race/ethnicity, B) age group among females, and C) age group among males. The black lines represent the timing of the four executive orders assessed in the analyses (US declares public health emergency, North Carolina statewide Stay-at-Home order, statewide Phase 2: Safer-at-Home order, and statewide Phase 2.5: Safer-at-Home order); grey lines represent the time of the other COVID-related executive orders.

The largest drop in unintentional MVC hospitalization rates was seen among older women (≥65 years old, average monthly change -6.00 hospitalizations per 1,000,000 female residents) after the U.S. declared a public health emergency, although decreases were seen among all females (average monthly change -0.93 to -3.93), Figure 3B. Overall, unintentional MVC hospitalization rates returned to 2019 levels within a few months among all females. Among males, adults 18-44 years old experienced almost no decrease in unintentional MVC hospitalization rates after the US declared a public health emergency, yet still saw a large spike in hospitalization rates after the Stay-at-Home order was issued (average monthly change -0.42 and +7.41 per 1,000,000, respectively), Figure 3C. Unintentional MVC hospitalization rates remained elevated among 18-44 year-old males until then end of 2020. Among 45-64 year-old males, a substantial drop (average monthly change -4.49 per 1,000,000) then rapid recovery to 2019 levels by the start of Phase 2: Safer-at-Home was seen.

### Disparities in unintentional non-MVC hospitalization rates during COVID-19

While White residents experienced higher hospitalization rates of non-MVC unintentional injuries prior to the COVID-19 pandemic (average 2019 rates: 47.5 per 1,000,000 White residents compared to 16.2-23.5 hospitalizations per 1,000,000 Black/African American, Hispanic/Latino, and other race residents), similar changes in hospitalization rates were seen across all racial/ethnic groups, Figure 4A. Similar to unintentional MVC hospitalizations, unintentional non-MVC hospitalizations rates dropped after the U.S. declared a public health emergency and returned to 2019 levels by the time major restrictions were first lifted (Phase 2: Safer-at-Home) among all groups.

**Figure 4.**
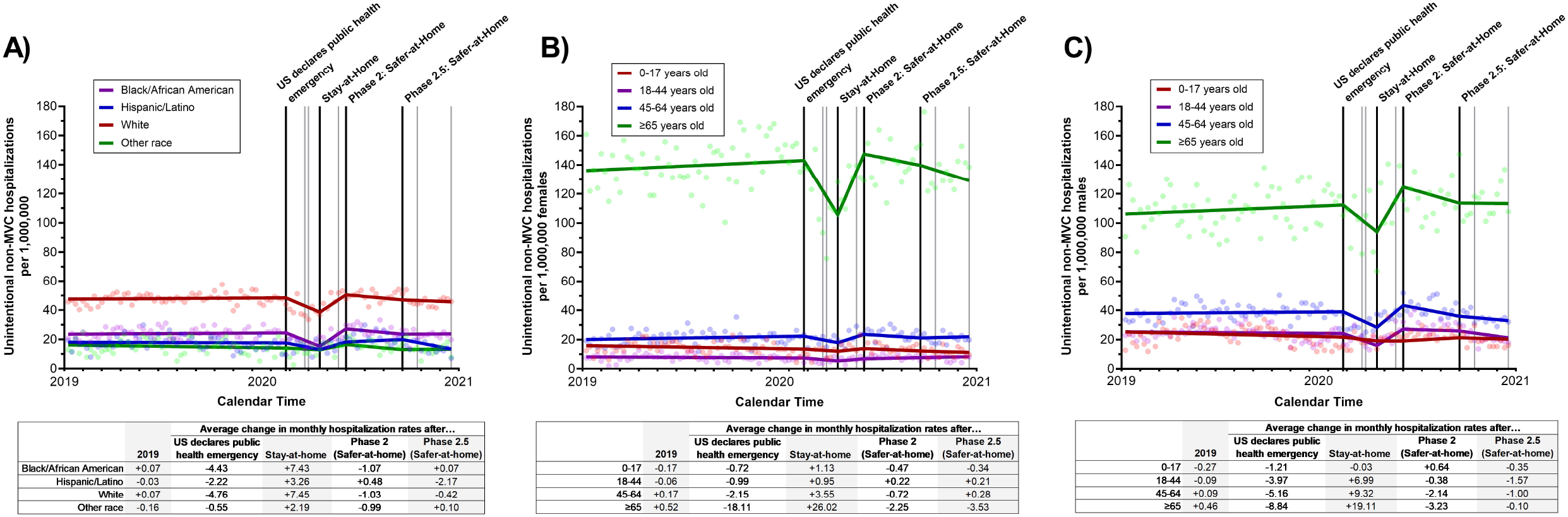
Impact of COVID-19 executive orders on weekly number of unintentional non-MVC admissions to trauma centers between January 2019 and December 2020 in North Carolina, stratified by A) race/ethnicity, B) age group among females, and C) age group among males. The black lines represent the timing of the four executive orders assessed in the analyses (US declares public health emergency, North Carolina statewide Stay-at-Home order, statewide Phase 2: Safer-at-Home order, and statewide Phase 2.5: Safer-at-Home order); grey lines represent the time of the other COVID-related executive orders.

Both older females and males (≥65 years old) experienced substantially higher rates of unintentional non-MVC hospitalizations (136.0 and 106.1 hospitalizations per 1,000,000 in 2019) compared to other age groups, Figures 4B and 4C. Among females 0-64 years-old, only modest declines were seen after the U.S. declared a public health emergency (average monthly change -0.72 to -2.15) compared to older females (average monthly change -18.11); a similar trend was seen among males. Unintentional non-MVC hospitalization rates bounced back to 2019 levels for all age groups (female and male) by the time major restrictions were first lifted.

## Discussion

In a statewide analysis of trauma hospitalizations, we found that the COVID-related policies were associated with changes in assault, self-inflicted, and unintentional injury hospitalization rates. When the statewide Stay-at-Home order was issued in North Carolina, assault hospitalization rates, primarily among Black/African American residents and adults aged 18-44, increased quickly but then dropped back to 2019 levels once restrictions had been lifted. After the U.S. declared a public health emergency, both unintentional MVC and non-MVC hospitalization rates decreased across most age groups, with the most substantial changes occurring in older adults. Interestingly, men aged 18-44 saw no declines in non-MVC injury hospitalizations after the declaration but still saw the same increase a few months later, with rates not falling to 2019 levels until the end of 2020. To the best of our knowledge, this is the first in-depth assessment of changes and disparities in trauma hospitalizations due to a statewide Stay-at-Home order and other COVID-related policies in the United States across race/ethnicity, age, and sex during the pandemic.

Increases in assault hospitalization rates, particularly firearm injuries, during the North Carolina statewide Stay-at-Home orders during the COVID-19 pandemic have been observed in other states in the U.S.(14, 15, 30). In our analysis, we also found that intentional injury hospitalization rates only increased among Black/African American residents, and adults aged 18-44 years old. The disparate effect of Stay-at-Home orders among Black/African American residents, compared to other racial/ethnic groups, may be partially explained by the increased burden of COVID-related financial, mental, and emotional strain(21, 22) among a population also at higher risk for experiencing assault. The increased rate of assault hospitalizations among women aged 18-44 indicate that statewide Stay-at-Home orders may have led to an increase in domestic violence, which was both a noted concern(31) and has been observed in other studies(17, 18). While we did not observe an increase in assaults among children, increases in child abuse have been observed in at least one U.S.-based study(16); it is possible that we were unable to detect a change due to the low baseline rate of assault hospitalizations among this age group. Overall, both our findings and those of other studies suggest that Stay-at-Home orders and other COVID-related policies in the U.S., and potentially other countries, had unintended consequences and that these were felt more among racial/ethnic minorities, women, and children.

The temporary decreases we saw at the beginning of the pandemic in unintentional injury (MVC and non-MVC) hospitalizations have also been observed in other U.S. states(6-9) and globally(32). Even prior to Stay-at-Home orders in the U.S.(3), many people began teleworking in the early months of the COVID-19 pandemic(33, 34), leading to fewer people commuting and fewer MVCs(8-10, 35), and likely fewer workplace injuries (unintentional non-MVCs). School closures, which occurred prior to the Stay-at-Home order in North Carolina, may have also led to decreased unintentional hospitalization rates early in the pandemic.

Interestingly, we observed no initial change in the unintentional MVC hospitalization rate among males 18-44 years-old, which is different from the trends we observed in every other sex/age group and inconsistent with previously reported findings of both fewer cars on the road(33-35) and fewer MVCs(6, 8-10) overall during the first several months of the pandemic in the U.S.(2) and abroad(1). And while several subgroups in North Carolina saw increases in unintentional MVC hospitalizations after these initial decreases, most stopped once hospitalization rates returned to pre-pandemic levels; among Black/African American residents and men aged 18-44 years old, assault hospitalization rates rose above 2019 levels and did not return to baseline until the end of 2020. Research is needed to identify potential behavior changes and policy effects that led to these prolonged increases in MVC hospitalizations among these individuals.

We also did not expect to see such substantial declines in unintentional non-MVC injury hospitalizations among older adults – the majority of which were fall-related – in the first few months of the pandemic. While one other U.S.-based study also stratified by age(6), they found no change in unintentional non-MVC hospitalizations among adults ≥65 years old. However, hospitals in NCTR have an older patient population, compared to that study (37.1% vs. 16.6% ≥65 years old in 2020), and we also assessed weekly rates and allowed trends to change across several COVID-related policies, instead of averaging across the entire COVID-period, which may explain the differences in our findings. This rapid decrease in non-MVC hospitalization rates in North Carolina suggest that older adults may have been avoiding seeking care at a hospital during the pandemic due to fear of COVID-19 infection(36). Declines in acute myocardial infarction and stroke hospitalizations (overall and among older adults) during the first few months of the COVID-19 pandemic, illnesses which should have not been impacted by the pandemic or Stay-at-Home orders, have also been reported in the United States(37, 38) and Europe(39, 40), further suggesting that people have avoided going to the hospital for necessary medical care. Policies and messaging are needed to ensure individuals seek needed urgent care for trauma during outbreaks and pandemics to avoid possible long-term, detrimental effects.

This study has a few limitations. First, the NCTR only includes individuals hospitalized for a traumatic injuries and therefore only captured a proportion of all traumatic injuries; individuals who were treated in an emergency department (but never admitted) or did not seek care at all would be missed. Given the known disparities in access to care(41) and trauma(19, 20) in the U.S., it is likely that we have underestimated the burden of serious traumatic injuries among racial/ethnic minorities. Similarly, only individuals who survived their initial injuries would be sent to a hospital for treatment and could also lead to unequal underestimation; however, we compared trauma rates within racial/ethnic and age/sex groups, which would minimize the effect of underestimation. Third, outside of the COVID-19 pandemic and related policies, there were several, co-occurring nationally recognized events that could also potentially have impacted trauma hospitalization rates. Thus, our results should be interpreted with some caution. Finally, self-inflicted injury hospitalization rates were too rare to conduct stratified analyses; future studies should utilize databases that capture both emergency department visits and deaths to elucidate concerns regarding self-inflicted injuries and suicide during COVID-19.

## Conclusions

Overall, it appears the Stay-at-Home orders implemented during the COVID-19 pandemic has had unintended consequences that disproportionately impacted racial/ethnic minorities and other marginalized groups in North Carolina, and potentially the United States. Fear of COVID-19 may have also led to decreases in unintentional non-MVC hospitalization rates, particularly among older adults, which may have long-term consequences. Given the potential far-reaching adverse impacts of national and statewide policies on racial/ethnic minorities and other high-risk groups, it is crucial for policy makers to anticipate possible negative effects and develop tailored, culturally appropriate approaches to mitigate harms that may disproportionately affect already disadvantaged communities.

## Data Availability

Data is available for request from the North Carolina Office of Emergency Medical Services. Researchers will make code available upon request. Please contact Dr. Paula Strassle for access.

## List of abbreviations

ED: emergency department
MVC: Motor vehicle collision
NCTR: North Carolina Trauma Registry
NCOEMS: North Carolina Office of Emergency Medical Services
U.S.: United States
SD: standard deviation
IRD: Incident rate difference
IRR: incident rate ratio

## Declarations

### Ethics approval and consent to participate

This study was deemed exempt by Institutional Review Board review (IRB protocols: 20-2117 and 000330).

### Consent for publication

Not applicable.

### Author’s contributions

Paula Strassle, Alan Kinlaw, and Sharon Schiro designed the study. Paula Strassle analyzed the data. Paula Strassle and Jamie Ko wrote the first draft. All of the authors critically reviewed and approved the final manuscript.

## Acknowledgements

The authors gratefully acknowledge the efforts of the North Carolina Office of Emergency Medical Services as well as the North Carolina Trauma Registry Hospitals.

